# A Remote Digital Memory Composite to Detect Cognitive Impairment in Memory Clinic Samples in Unsupervised Settings using Mobile Devices

**DOI:** 10.1101/2021.11.12.21266226

**Authors:** David Berron, Wenzel Glanz, Lindsay Clark, Kristin Basche, Xenia Grande, Jeremie Güsten, Ornella V. Billette, Ina Hempen, Muhammad Hashim Naveed, Nadine Diersch, Michaela Butryn, Annika Spottke, Katharina Buerger, Robert Perneczky, Anja Schneider, Stefan Teipel, Jens Wiltfang, Sterling Johnson, Michael Wagner, Frank Jessen, Emrah Düzel, the DELCODE Consortium

**Affiliations:** German Center for Neurodegenerative Diseases, Magdeburg, Germany; Institute for Cognitive Neurology and Dementia Research, Otto-von-Guericke University, Magdeburg, Germany; German Center for Neurodegenerative Diseases, Bonn, Germany; University Department of Neurology, Medical Faculty University Hospital, Magdeburg, Germany; German Center for Neurodegenerative Diseases, Munich, Germany; Institute for Stroke and Dementia Research (ISD), University Hospital, LMU Munich, Munich, Germany; Department of Psychiatry and Psychotherapy, University Hospital, LMU Munich, Munich, Germany; German Center for Neurodegenerative Diseases, Tübingen, Germany; Section for Dementia Research, Hertie Institute for Clinical Brain Research and Department of Psychiatry and Psychotherapy, University of Tübingen, Tübingen, Germany; Department for Neurodegenerative Diseases and Geriatric Psychiatry, University Hospital Bonn, Bonn, Germany; Department of Psychosomatic Medicine, Rostock University Medical Center, Rostock, Germany; German Center for Neurodegenerative Diseases, Rostock, Germany; German Center for Neurodegenerative Diseases, Göttingen, Germany; Department of Psychiatry and Psychotherapy, University Medical Center Göttingen, University of Göttingen, Göttingen, Germany; German Center for Neurodegenerative Diseases, Cologne, Germany; neotiv GmbH, Magdeburg, Germany; Munich Cluster for Systems Neurology (SyNergy), Munich, Germany; Ageing Epidemiology Research Unit (AGE), Imperial College London, London, UK; Department of Medicine, Division of Geriatrics and Gerontology, University of Wisconsin, School of Medicine and Public Health, Madison, Wisconsin, US; Geriatric Research Education and Clinical Center, William S. Middleton Memorial Veterans Hospital, Madison, Wisconsin, US

**Keywords:** digital cognitive assessment, episodic memory, subjective cognitive decline (SCD), mild cognitive impairment (MCI), Alzheimer’s Disease (AD)

## Abstract

Mobile app-based unsupervised monitoring of cognition holds the promise to facilitate case-finding in clinical care and the individual detection of cognitive impairment in clinical and research settings. In the context of Alzheimer’s disease, this is particularly relevant for patients who seek medical advice due to memory complaints. Here we developed a Remote Digital Memory Composite (RDMC) score from an unsupervised remote and mobile cognitive assessment battery focused on episodic memory and long-term recall and assessed its construct validity, retest reliability and diagnostic accuracy when predicting MCI-grade impairment in a memory clinic sample and healthy controls. A total of 199 study participants were recruited from three cohorts and included as healthy controls (HC; n=97), individuals with subjective cognitive decline (SCD; n= 59) or patients with mild cognitive impairment (MCI; n=43). Study participants performed cognitive assessments in a fully remote and unsupervised setting via a smartphone app for cognitive testing. The derived RDMC score was highly correlated with the PACC5 score across participants and demonstrated good retest reliability. Diagnostic accuracy for discriminating memory impairment from no impairment was high (cross-validated AUC = 0.83, 95% CI [0.66, 0.99]) with a sensitivity of 0.82 and a specificity of 0.71. Our results indicate that unsupervised mobile cognitive assessments in a memory clinic setting using the implementation in the neotiv digital platform results in a good discrimination between cognitively impaired and unimpaired individuals. Thus, it is feasible to complement neuropsychological assessment of episodic memory with unsupervised and remote assessments on mobile devices. This contributes to recent efforts for implementing remotely performed episodic memory assessment for case-finding and monitoring in large research trials and clinical care.

## Background

Differentiating mild cognitive impairment (MCI) from subjective cognitive impairment is important to provide prognosis regarding future cognitive decline as well as regarding the potential eligibility for treatments at the MCI stage of Alzheimer’s disease (AD). However, differentiating MCI from subjective cognitive impairment is still very challenging using brief cognitive tests ^1^. More than 80% of older adults who seek medical advice due to memory complaints and who are later found to have a biomarker profile indicative of Alzheimer’s disease have an amnestic variant with predominant episodic memory impairment ^2^. Indeed, episodic memory, the ability to recall spatial and temporal relationships of personally experienced events ^3^, is a key component of the neuropsychological assessment of individuals with suspected AD ^4^. Consequently, episodic recall is an important element of the Preclinical Alzheimer Cognitive Composite (PACC5) ^5,6^.

The aim of the PACC5 is to provide a comprehensive assessment of AD relevant cognitive impairment and to serve as a tool with validated sensitivity to detect cognitive decline over time ^5,6^. However, traditional paper-and-pencil assessments to derive composites such as the PACC5 are time-consuming and require supervision by a trained neuropsychologist ^6^. This severely restricts their utility and implementation in primary care, especially when considering equal-opportunities to comprehensive neuropsychological assessments also in rural areas, and use of high frequency cognitive assessment to account for day-to-day variations in cognitive performance. In general, the long test duration and specialized supervision make the high-frequency longitudinal use of established neuropsychological assessments practically impossible. There is, thus, a strong need for unsupervised, remote, high-frequency cognitive assessment that can provide meaningful approximation of traditional neuropsychological scores.

Mobile devices such as smartphones and tablets have the potential to enable unsupervised and remote high-frequency cognitive assessments ^7–9^. Indeed, recent studies have demonstrated the feasibility of remote digital cognitive assessments in the general population ^10^ as well as in older adults and patient populations ^11–16^. Implementing a mobile and remote proxy for a neuropsychological assessment (such as the tests that comprise the PACC5) also offers the opportunity to overcome some of the shortcomings of neuropsychological tests. One potential disadvantage of established neuropsychological assessments of episodic memory is for example that they heavily tax on verbal abilities which makes it difficult to assess episodic memory in multi-lingual settings or when verbal abilities are already impaired ^4^. In addition, implementing new cognitive tests allows to take into account the latest insights into the functional architecture of episodic memory and the spread of AD pathology. Recent work on the functional neuroanatomy of episodic memory showed that episodic memory involves a network including medial temporal, midline parietal and cortical regions, each of which serve different functions and are affected in different stages of AD ^17,18^. Episodic memory requires pattern separation processes that are mediated by the dentate gyrus ^19,20^ and reduce memory interference between similar events, and pattern completion processes that are mediated by hippocampal Cornu Ammonis 3 (CA3) and enable the recollection of details from a past event in interplay with neocortical regions ^21^. The medial temporal lobe regions provide information to the hippocampus mainly through the entorhinal cortex. That in turn, receives partly domain-segregated information such that object representations are transferred via the perirhinal cortex and the anterior-lateral entorhinal subdivision and scene representations via the parahippocampal cortex and posterior-medial parts of the entorhinal cortex ^22–26^. Taken together, there is converging evidence that in addition to long-term recall, short-term mnemonic discrimination of object and scene representations is impaired in the predementia stages of AD ^27^. Besides pattern separation and completion, a third aspect of episodic memory is recognition memory ^28,29^. Although the neurobiology of recognition memory is complex and it is likely to have a non-episodic, familiarity-based component ^30–32^, it is evident that medial temporal lobe dysfunction can impair recognition memory alongside impairments of recall ^30^.

A set of anatomically informed and non-verbal tasks for episodic memory that incorporate these recent insights into the functional anatomy of episodic memory is available on the neotiv digital platform (https://www.neotiv.com/en) ^10,15^ and has been implemented in prospective cohort studies of the German Center for Neurodegenerative Diseases (DZNE) and the Wisconsin Registry for Alzheimer’s Prevention (WRAP). There are three different tests of memory. First, a short-term mnemonic discrimination test tapping into pattern separation, separately implemented for object and scene stimuli, second, a short- and long-term cued-recall test of object-scene associations tapping into pattern-completion and, third, a long-term photographic scene recognition memory test.

Here we evaluated these three memory measures in a remote and unsupervised design using mobile devices. To that end, we developed a Remote Digital Memory Composite (RDMC) score and assessed its construct validity using in-clinic neuropsychological assessment as well as its retest reliability across independent test sessions. Finally, we assessed the diagnostic accuracy of the RDMC score when differentiating between individuals with and without cognitive impairment in a memory clinic sample.

## Materials and Methods

### Participants and neuropsychological assessments

DELCODE is an observational longitudinal memory clinic-based multicenter study in Germany, with participants aged 60 years and older, that either have mild dementia of the Alzheimer’s type, amnestic mild cognitive impairment (MCI), subjective cognitive decline (SCD), or are asymptomatic healthy controls (HC). The detailed study design of DELCODE is reported in ^33^. The remote mobile monitoring add-on study started in April 2019 after a separate approval by the ethical committees of each participating site. The annual neuropsychological testing in DELCODE included the PACC5 ^5^ and other assessments reported in full in ^33^. The PACC5 z-score was calculated as the mean performance z-score across the MMSE ^34^, a 30 item cognitive screening test, the WMS-R Logical Memory Delayed Recall ^35^, a test of delayed (30 min) story recall, the Digit-Symbol Coding Test (DSCT; 0–93) ^36^, a test of memory, executive function and processing speed, the Free and Cued Selective Reminding Test–Free Total Recall (FCSRT96; 0–96) ^37^, a test of free and cued recall of newly learned associations, and the Category Fluency Test, a test of semantic memory and executive function. The z-scores for the PACC5 elements were derived using the mean and standard deviation of HC and SCD from the baseline visit of the entire DELCODE study sample. A PACC5 composite score was calculated when at least three of its five components were available while making sure that at least the MMSE, one memory measure and either category fluency or DSCT were included. In the DELCODE cohort, the clinical labels (HC, SCD, MCI) were established at the baseline assessment of each participant and follow-up diagnoses have been established in April 2021 (Stark et al., submitted). At the Memory Clinic of the University of Magdeburg, which is a DELCODE recruitment site, additional participants were prospectively recruited according to the DELCODE neuropsychological protocol. Recruitment criterion for these additional participants was clinically suspected MCI on the basis of progressive memory complaints. WRAP is a longitudinal observational cohort study enriched with persons with a parental history of probable AD dementia ^38^. The neotiv add-on study began in April 2021. Neuropsychological testing in WRAP included the PACC5 ^5^ and was completed every two years. The PACC5 z-score was calculated as the mean performance z-score across the MMSE ^34^, a 30 item cognitive screening test, the WMS-R Logical Memory Delayed Recall ^35^, a test of delayed (30 min) story recall, the Digit-Symbol Coding Test (DSCT; 0–93) ^36^, a test of memory, executive function and processing speed, the Rey Auditory Verbal Learning Test Total Learning Trials (RAVLT-Total; 0-75)^49^, a test of verbal learning, and the Category Fluency Test (animals in WRAP, animals and food in DELCODE) a test of semantic memory and executive function. The z-scores for the PACC5 elements were derived using the mean and standard deviation of cognitively unimpaired participants from the baseline visit of the WRAP cohort.

### Study design

All participants except patients with dementia were eligible if they owned a smartphone or tablet with internet access that was technically suitable for the mobile app to be installed on and that they could operate on their own. In addition, participants with sensory or motor problems or with significant neurological illness that may interfere with the ability to complete tasks were not eligible. DELCODE participants were asked at their regularly scheduled annual follow-up visit and memory clinic patients during their in-clinic visit whether they would like to participate in the add-on study. If they agreed, study personnel did lend support installing the app from the respective app store on the participants’ own mobile device, but participants received no further verbal instructions apart from a printed manual. In WRAP, those who met eligibility criteria were sent a recruitment letter and the consent form and were contacted over the telephone ∼2 weeks thereafter and invited to participate. If interested, a telephone screening was completed to confirm eligibility. Following screening, the informed consent process was completed and verbal consent was obtained from the participant. Instructions for how to download the app were emailed, and followed up over telephone to provide additional assistance on downloading the app if needed.

The Object-in-Room Recall test (ORR), the Mnemonic Discrimination Test for Objects and Scenes (MDT-OS) and the Complex Scene Recognition Test (CSR) were completed by participants remotely and unsupervised using their mobile device. In all cohorts, participants were asked to complete memory assessments every two weeks. In WRAP, there was also an additional subgroup of participants who were asked to complete a burst of the same number of memory tasks every other month over four consecutive days. Each assessment consisted of a 2-phase session separated by a short delay. The two phases were either two halves of mnemonic discrimination, or encoding and retrieval phases of complex scene recognition and object-in-room recall (see details of the tasks below). Every phase took around 5-10 minutes. Tests were remotely initiated via push notifications which were sent at the same time-of-day as the registration, but participants had the possibility to postpone test sessions. This approach was chosen not to urge participants to take the test under suboptimal conditions such as distraction, fatigue or temporary illness. Daily reminders were sent via push notifications until the respective task was completed, and the actual time of testing was recorded. Before each test session, participants were reminded by the app to perform the test in a quiet environment, to put their glasses on if needed and to ensure that their screen was bright enough to see the pictures clearly. They also received a short practice session at the beginning of each session. After each test session, participants were asked within the app if they were distracted by things happening around them during the session (yes/no decision) and to rate their concentration level and subjective performance (1=very bad, 2=bad, 3=middling, 4=good and 5=very good). Hence, participants received the instructions for the cognitive tests remotely and performed the test fully unsupervised.

#### Mnemonic Discrimination Test of Objects and Scenes (MDT-OS)

Figure 1A shows the outline of the MDT-OS test ^22,23,25,39^. In this test, participants are presented with 3D rendered computer-generated objects and scenes that are repeated either identically or in slightly modified versions. Participants need to decide whether a repeated presentation shows a repetition of the original picture or a modified version. They indicate their response by either tapping on a button (for an exact repetition) or by tapping on the location of a change (for a modified version). They see 32 object and 32 scene pairs where half are repeated or modified respectively. One session was split into two phases and completed on two different days. The first phase was presented as a one-back task while the second phase was presented as a two-back task. The test provides a hit rate, a false alarm rate and a corrected hit rate for both the object and scene condition. The averaged corrected hit rate across the scene and the object condition is used for the RDMC.

**Figure 1:**
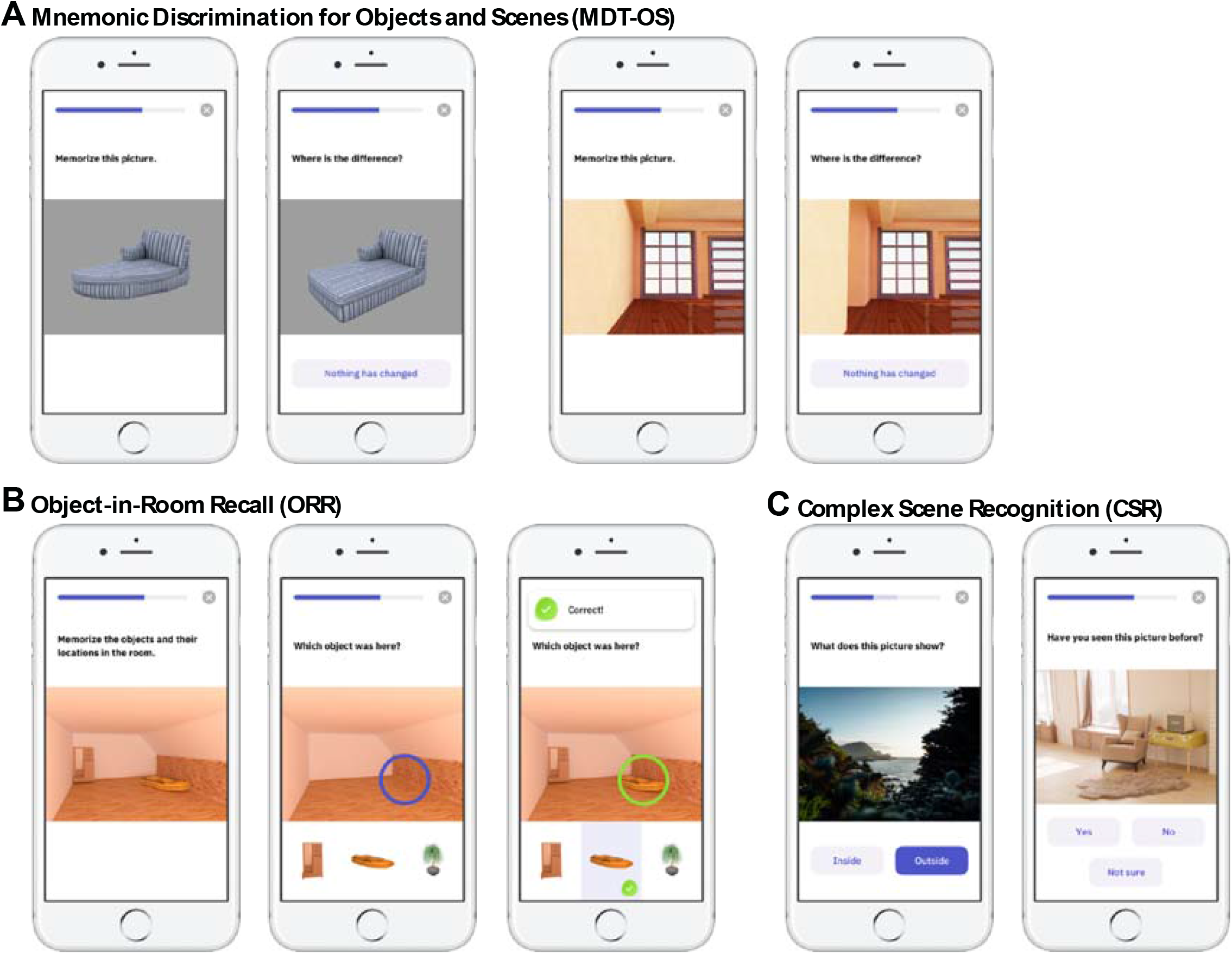
Memory tests constituting the RDMC score. Mnemonic Discrimination Test for Objects and Scenes (MDT-OS), Object-in-Room-Recall (ORR) test and Complex Scene Recognition Test (CSR).

#### Object-in-Room Recall Test (ORR)

Figure 1B shows the outline of the ORR-Test (for a discussion of the principles of pattern completion on which this test is based see ^21^). In this test, participants were asked to memorize 3D rendered computer-generated rooms, in which two 3D-rendered objects are placed. Participants recall which object was placed at a specific location cued by a colored circle in the empty room in an immediate recall test. They indicate their recall decision by tapping on one of three objects displayed below the empty room: the correct object for that location, the object that was also present in the room but at a different location (correct source distractor) and a completely unrelated object (incorrect source distractor). They learn 25 such object-scene associations. After a delay of 30 minutes (DELCODE and Memory Clinic) or 90 minutes (WRAP), participants are prompted to complete a delayed recall test. However, as can be seen in the results section, the actual delay period based on the real time of test completion was similar across cohorts. In the ORR test, the ability to recall the correct association is graded and allows to separate correct episodic recall from incorrect source memory. Thus, correct recall excludes the choice of an object that was present in the same room but at a different location (wrong source memory for specific location) and an object that was not present in the room but nevertheless associated with the objects belonging to the room during encoding (wrong source memory for overall location). The test provides an immediate (0-25) and a delayed recall score (0-25). For the RDMC we use the total recall, which is calculated as the average of the z-standardized immediate and delayed recall score.

#### Complex Scene Recognition Test (CSR)

Figure 1C shows the outline of the CSR test ^28,29,40^. Participants see 60 photographic images depicting indoor and outdoor scenes. For encoding, participants make a button-press decision whether the presented scene is indoors or outdoors. After an instructed delay of 65 (DELCODE) or 90 (WRAP) minutes, the participants are informed via push notification to complete the second phase of the task. Here, the encoded images are presented together with 30 new images and participants make old/new/uncertain recognition memory decisions. The test provides a hit rate, a false alarm rate and a corrected hit rate. The corrected hit rate is used for the RDMC.

### Data handling

Participants used the app with a pseudonymized ID (no identifying information or clinical information was available or required in the mobile app) provided to them during an in-clinic visit or over the telephone. The app data were transferred to the research centers in accordance with the General Data Protection Regulation. The mobile app data were then related to the clinical data by the research centers and in the following released to Principal Investigators and to neotiv GmbH. Data handling and quality control procedures for the clinical DELCODE data are reported in ^33^.

### Quality Control Procedures

For this manuscript, we analyzed data up until the data release on July 7^th^ 2022 in DELCODE and March 3^rd^ 2022 in WRAP. Recruitment of participants in the Memory Clinic Magdeburg was performed between October 2020 and January 2022. Regarding the sessions from the first wave, meaning each very first assessment of CSR, MDT-OS and ORR, 8% of test sessions exceeded the threshold for missing responses (maximum of 20%) and 26.5% of test sessions exceeded the maximum length of the delay period (180 minutes) before filtering. Thus, these sessions were excluded during quality assessment. Excluded test sessions were replaced by valid subsequent sessions where possible. As a result, 82% of test sessions we report here were from the first wave and 18% from subsequent test sessions.

### Statistical analysis

All statistical analyses were performed in R (R Core Team, 2020). We correlated the RDMC score with in-clinic neuropsychological assessments to assess construct validity using the Pearson correlation coefficient. Multiple regression models were used to assess the relationship with age, sex and years of education on the PACC5 as well as the RDMC. We conducted these analyses within the sample from the DELCODE study and Memory Clinic of the University of Magdeburg, given that a PACC5 score with slightly different elements was used in the WRAP cohort. In addition, we assessed the influence of the Time-to-Retrieval and the screen size of the mobile device on the individual components of the RDMC as well as on the RDMC itself. We also assessed retest reliability using the Pearson Correlation Coefficient and the Intraclass Correlation Coefficient. Diagnostic accuracy and receiver operating characteristic (ROC) analyses were performed using the pROC package ^41^. A cross-validated Area under the Curve (cvAUC) was derived from 5-fold cross-validation. Logistic regression models including participant age, sex and years of education were used to compare models containing the RDMC to models only containing demographic information.

### Data availability

Data, study protocol and biomaterials can be shared with partners based on individual data and biomaterial transfer agreements.

## Results

### Participant characteristics and recruitment

Here we considered the first 199 study participants who completed at least one session of each of the three cognitive tests (97 HC, 59 SCD, and 43 MCI). Following quality assurance as described above, 143 individuals could be included who contributed at least one valid test session per paradigm.

We calculated a PACC5 cut-off score across all non-demented participants in the DELCODE and WRAP cohorts baseline assessment, respectively, that distinguished MCI from cognitively unimpaired participants (HC and SCD) prioritising sensitivity > 0.8, and resulting in a cut-off of -0.515 for DELCODE and -0.981 for WRAP. In order to make sure to use the latest information on participant’s cognitive status, cognitive impairment was then defined as either an MCI diagnosis (at the baseline or follow-up assessment) or a PACC5 score suggestive of mild cognitive impairment at the closest follow-up session to the app-based assessment. This resulted in cognitively unimpaired (CU n=105) and cognitively impaired participants (CI n=38; see table 1 for participants’ characteristics).

**Table 1:** Participant demographics of the entire sample

|  | HC<br>(N=78) | SCD<br>(N=40) | MCI<br>(N=25) | CU<br>(N=105) | CI<br>(N=38) | Total<br>(N=143) |
| --- | --- | --- | --- | --- | --- | --- |
| Age (years) | 68.2 (5.47) | 69.2 (7.67) | 69.2 (6.79) | 67.9 (6.01) | 70.7 (6.92) | 68.6 (6.36) |
| Education (years) | 15.9 (2.82) | 14.6 (2.64) | 13.9 (2.49) | 15.7 (2.79) | 13.9 (2.55) | 15.2 (2.82) |
| Sex (female) | 67.9 % | 52.5 % | 36 % | 62.9 % | 44.7 % | 58 % |
| MMSE | 29.5 (0.72) | 29.4 (0.9) | 26.9 (2.74) | 29.6 (0.69) | 27.5 (2.41) | 29 (1.64) |
| RDMC | 0.01 (0.67) | -0.21 (0.6) | -1.03 (0.69) | 0 (0.65) | -0.87 (0.66) | -0.23 (0.76) |

*\*displays mean values (standard deviations) unless otherwise stated*
*\*Abbreviations: N, number of participants; HC, Healthy Controls; SCD, subjective cognitive decline; MCI, mild cognitive impairment; MMSE, mini-mental state examination; CU and CI, cognitively (un)impaired based on clinical diagnosis or the most recent PACC5.*

### Contextual factors

Across all three cognitive tests, participants reported high concentration levels during the task (mean = 3.99, scale 1-5, which translates to good concentration), and moderately high subjectively rated task performance (mean = 3.51, scale 1-5 which translates to middling subjectively rated performance). While concentration levels were similar across tasks (3.64, 4.08, 4.23 for MDT-OS, ORR and CSR respectively), subjective performance indicated higher task difficulty for the MDT-OS (2.9) compared to ORR and CSR (3.66 and 3.97 respectively). In addition, participants reported no distractions during their test sessions in 92% of the tasks (MDT-OS: 90%, ORR: 92%, CSR: 96%). Following filtering, the time between encoding and retrieval in the ORR and CSR tests was on average 67 minutes (SD = 36) and 92 minutes (SD = 23) respectively. In DELCODE, participants were invited to the retrieval phase of the ORR after 30 minutes, and their actual mean delay was 49 minutes, while they were invited to the CSR retrieval after 65 minutes, and completed it on average after a delay of 82 minutes. In WRAP, participants were invited to the retrieval phases of both paradigms after 90 minutes and completed it on average after 97 minutes and 101 minutes for the ORR and CSR task respectively. Mobile devices had a screen diagonal between 10.2 – 32.9 cm (mean 16 cm, SD = 5.5) indicating the use of smartphones as well as tablet computers.

### Development of the Remote Digital Memory Composite

We built the RDMC using equal weights where each component (each of the three cognitive tests) had the same weight. While the CSR only provides a corrected hit rate from the delayed recall, the ORR and the MDT provide two outcomes. In the ORR, there is an immediate as well as a delayed recall while the MDT comes in two task conditions, one corrected hit rate for scenes and one for objects ^10,15^. All individual components (ORR-Im, ORR-Del, MDT-O, MDT-S and CSR) were z-standardized using the mean and standard deviation of the cognitively unimpaired participants (CU). For the RDMC, we then calculated the z-scored mean across ORR-Im and ORR-Del (TotalRecall), the z-scored mean score across MDT-O, MDT-S (TotalCorrectedHitRate) and the z-scored corrected hit rate of the CSR. The resulting three z-scores were averaged to derive the final RDMC score. The retest reliability between two independent time points was good (r = 0.8, p<.001; ICC = 0.8).

### Relationship between the RDMC and the PACC5

In order to assess construct validity, we used neuropsychological data from the closest-in-time in-clinic visit in the DELCODE study (to the mobile app add-on study) and Memory Clinic to perform a correlation analysis between the RDMC and the PACC5 score as well as its individual elements. The average time interval between the in-clinic visits and the remote app assessments was 80 days. The first RDMC correlated highly (r=.62, p<.001) with the closest-in-time in-clinic PACC5 scores (see Figure **2**). When considering only participants with memory complaints, meaning those that were referred to the memory clinics by their general practitioner and fulfilled either SCD or MCI criteria, the construct validity of the RDMC remained high (r = .62, p<.001). The construct validity in individuals without memory complaints (HC) was similar (r = .59, p=.001). We used hierarchical multiple regression models to test whether each element of the RDMC (ORR-Total, MDT-OS and CSR) added significantly to predicting the PACC5 score. Model comparisons using the Akaike Information Criterion (AIC) indicated a significant effect for each predicting component. In addition, we assessed the relationship of the RDMC with each of the individual elements of the PACC5. The RDMC correlated strongest with the memory measures, the Logical Memory Delayed Recall (*r*=0.55, *p*<0.001) and the FCSRT96 (*r*=0.53, *p*<0.001) but also with the DSCT (*r*=0.54, *p*<0.001). As expected, we found weaker relationships with the MMSE (*r*=0.49, *p*<0.001) and verbal fluency (*r*=0.34, *p*=0.001).

**Figure 2:**
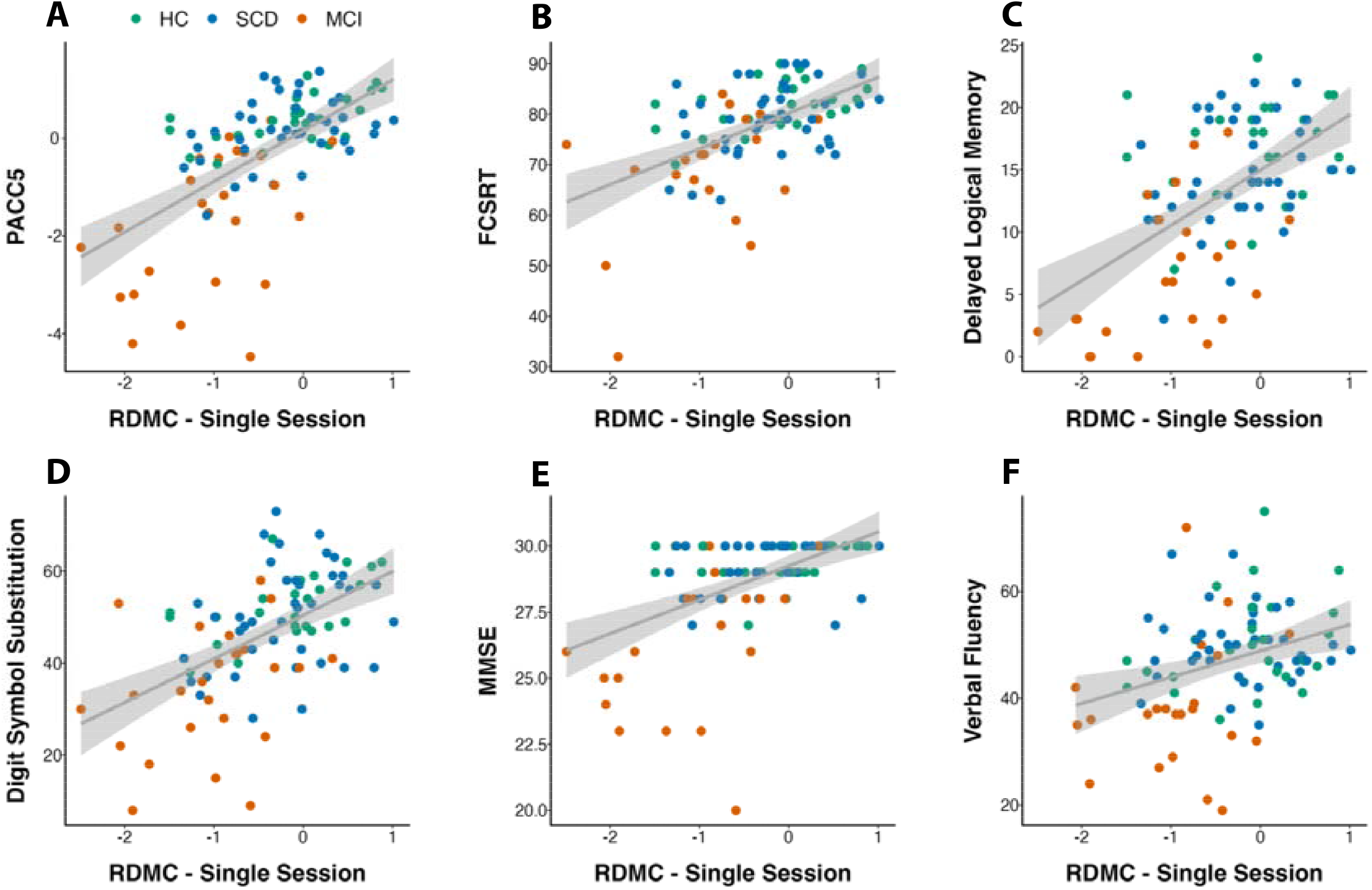
Construct validity of the composite score. Correlation between the RDMC (first test) and the closest-in-time (A) DELCODE PACC5 assessment as well as (B-F) all individual PACC5 elements. HC, Healthy Controls; SCD, subjective cognitive decline; MCI, mild cognitive impairment; FCSRT, Free and Cued Selective Reminding Test; MMSE, Mini Mental State Exam.

**Figure 3:**
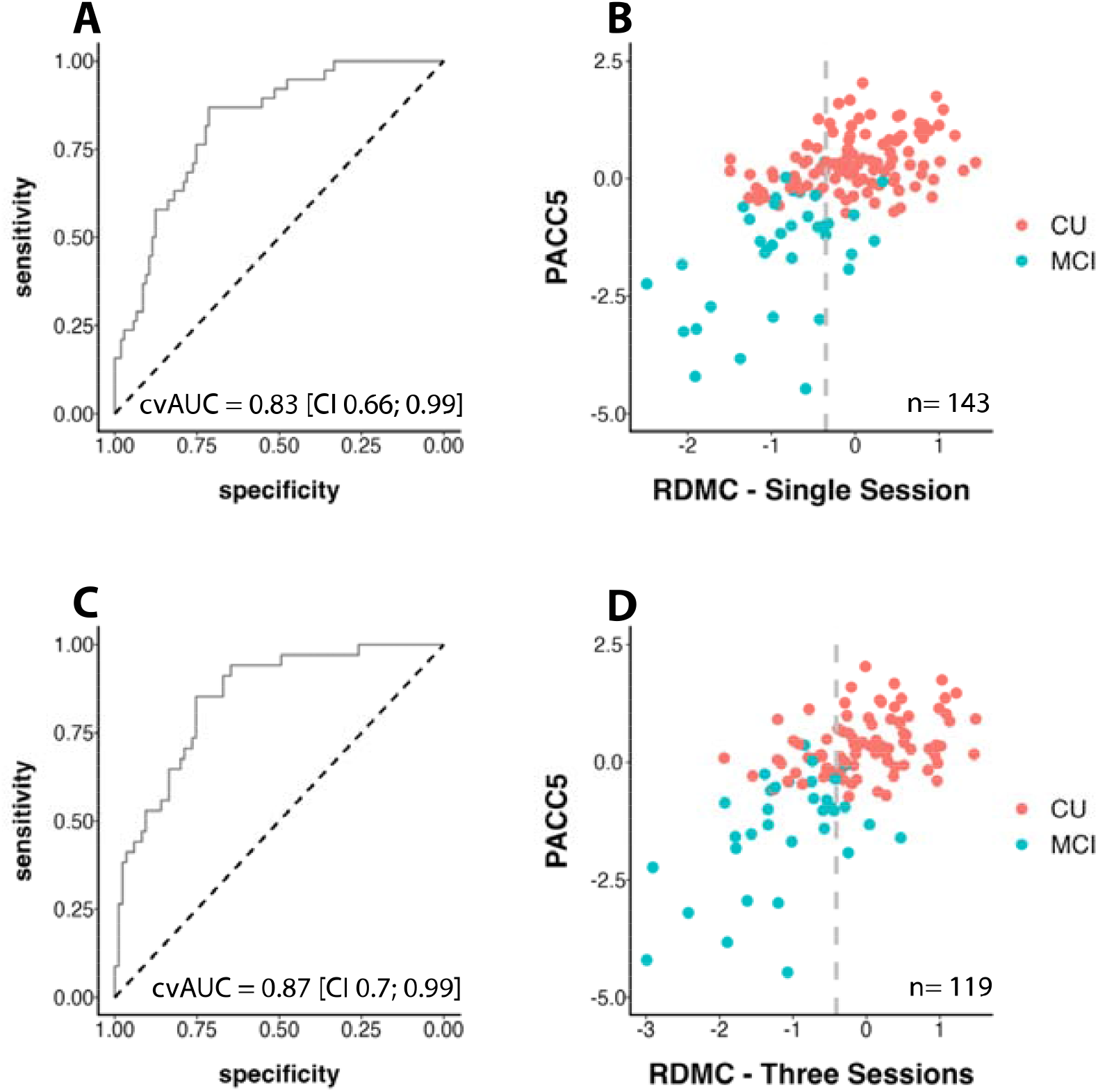
Diagnostic accuracy across Single Session and Three Session regimens. (A) and (C) Receiver Operating Characteristic (ROC) curve showing diagnostic accuracy for the detection of MCI-grade cognitive impairment based on the RDMC. (B) and (D) Scatter plot showing the optimal RDMC cut-off with CU and CI in different colors – optimal cut-offs at -0.35 and -0.41 indicated by the dashed grey line, dots to the left are classified as cognitively impaired, and dots to the right as cognitively unimpaired.

### Relationship with Age, Sex, Education and other factors

Multiple regression models with age, sex, years of education, time-to-retrieval and screen size were calculated to identify the relationships with individual components of the RDMC. For CSR, only years of education was a significant predictor where higher education was associated with better performance (β_education_ = 0.12, p<0.001; β_age_ = -0.007, p=0.6; β_sex_ = -0.29, p=0.1; β_delay_ = 0.19, p=0.38; β_screen_ = -0.003, p=0.839). For MDT-OS, only age was a significant predictor where higher age was associated with worse task performance (β_age_ = -0.027, p=0.02; β_sex_ = -0.19, p=0.204; β_education_ = 0.02, p=0.464; β_screen_ = -0.02, p=0.159). For the ORR, age and years of education were associated with task performance (β_age_ = -0.05, p<0.001; β_education_ = 0.06, p=0.049; β_sex_ = -0.1, p=0.531; β_delay_ = 0.21, p=0.139; β_screen_ = 0.009, p=0.518). With respect to the RDMC, more years of education (β_education_ = 0.09, p=0.002) as well as younger age (β_age_ = -0.04, p=0.002) were associated with higher task performance, but not sex. In comparison, the PACC5 was associated with sex (β_sex_ = -0.66, p<0.001), years of education (β_education_ = 0.09, p<0.001) and age (β_age_ = -0.03, p=0.03), i.e., women and participants with more years of education and those that were younger obtained a higher PACC5 score.

### Diagnostic accuracy

The RDMC score differentiated CI and CU individuals with a cross-validated Area under the Curve (cvAUC) of 0.83 [CI 0.66; 0.99] resulting in a sensitivity and specificity of 0.82 and 0.72 respectively (optimal cut-off = -0.35). The ROC curve and classifications with the cut-off are presented in Error! Reference source not found.A. Alternatively, considering only MCI patients or only participants with a PACC5 score below the MCI-grade cognitive impairment cut-off as cognitively impaired yielded very similar cvAUCs of 0.85 [CI 0.66; 0.99] and 0.84 [CI 0.67; 0.98] respectively. In order to test whether all three components of the RDMC are needed to achieve the best possible classification, we performed individual AUC analyses for each individual component (ORR = 0.78; MDT-OS = 0.78; CSR = 0.72) as well as for alternative composite scores covering all possible combinations of only two test paradigms (ORR/MDT-OS: = 0.82; ORR/CSR = 0.8; MDT-OS/CSR = 0.79). No individual component or composite combining two components could however reach an AUC of 0.83.

In addition, we used logistic regression models to test how a model containing the RDMC would compare to a model containing only participant age, sex and years of education. The model that included the RDMC as predictor (Tjur’s *R*^2^ = 0.325; AIC = 126.4) fit the data significantly better than the basic model that included participants age, sex, and years of education (Tjur’s *R*^2^ = 0.160; AIC = 151.4) and outperformed its diagnostic accuracy (AUC: 0.85 compared to AUC: 0.74). A maximum sensitivity of 84% could be achieved given at least 70% specificity, and a maximum specificity of 85% could be achieved given at least 70% sensitivity.

### Health Care Scenario

Regarding potential challenges in broader use within health care, we designed an additional analysis schedule that would improve reliability and robustness above and beyond the provision of a single test. For this purpose, we considered 4 repetitions of each test where the worst test performance of each paradigm is removed. This provides redundancy to account for distractions and interruptions that might appear and allows averaging across up to three test sessions to increase the representativeness of the overall test result. Therefore, we only included participants who provided up to 4 valid test sessions of each paradigm, removed each participant’s worst performance per paradigm, calculated a mean score for each outcome and the respective z-score regarding the mean and standard deviation of the CU group before we calculated a composite by averaging across the 3 resulting means. For this analysis, 119 participants (85 CU and 34 CI) could be included. In this scenario, the RDMC score differentiated cognitively impaired (CI) and cognitively unimpaired individuals (CU) with a cross validated cvAUC of 0.87 [CI 0.7; 0.99] and a sensitivity and specificity of 0.85 and 0.74 respectively (optimal cut-off = -0.41). The ROC curve and classifications with the cut-off are presented in **Error! Reference source not found**.C.

## Discussion

We developed an unsupervised, self-administered and remote digital memory composite based on a single test session from each of three equally weighted memory tests (ORR, MDT-OS and CSR), which were performed remotely and fully unsupervised. The resulting RDMC showed high construct validity in relation to the PACC5 score, especially with its elements measuring memory, and good retest reliability in a subsample that performed each test twice. Finally, the RDMC could differentiate between individuals with and without MCI-grade impairment with an AUC of 0.83 demonstrating high diagnostic accuracy. An additional regimen with up to three individual test sessions per memory test with the aim to provide redundancy to account for potential distractions or interruptions while self-administrating the tests yielded an AUC of 0.87.

In terms of construct validity, we found a strong correlation between the RDMC and the PACC5, as well as its individual elements. This correlation was present in both cognitively healthy older adults and those with cognitive impairment indicating that the correlation was not driven by collating an impaired and a non-impaired group as two extremes into the same analysis. The fact that the correlation also held within those with cognitive symptoms (SCD and MCI) and that all of these individuals were recruited on the basis of referrals (as opposed to recruitment advertisements) provides first evidence that may support construct validity in a health care setting. In terms of reliability, we found a high correlation between two different instances of the RDMC conducted within a time interval of ∼6 weeks.

The RDMC identified individuals with an MCI-grade impairment with an AUC between 0.83 (single self-administration of each test) and 0.87 (three self-administrations of each test). This allowed to identify individuals with MCI-grade impairment with a sensitivity of 0.82 and a specificity of 0.72 on the basis of a single assessment of the RDMC. This is comparable or higher than several other recently reported unsupervised ^42^, or in-clinic and supervised digital cognitive assessments ^43–46^. Importantly, however, several earlier approaches reported outcomes by comparing MCI patients against samples that exclusively consisted of healthy asymptomatic older adults ^42–47^. In health care settings, the main challenge is to identify significant impairment within individuals with memory concerns. Therefore, we believe that our focus on memory complainers and the inclusion of a large number of SCD patients who sought medical advice (hence were not recruited through advertisements) in this sample is a major advantage in the validation and critical for future application.

Usability is a major limitation for mobile device-based assessments of cognition in old age and particularly in preclinical and prodromal AD. While participants were assisted during the installation of the neotiv mobile app and received a printed manual at the time when their consent was obtained at the memory clinic, all three tests were conducted fully remotely and without supervision. Participants received a push-notification on their mobile device each time a test was available to be performed. All instructions and guidance for performing the tests was provided in the app and included a training run of each test. Participants were also instructed to seek a quiet place where they would not be distracted and after each test were inquired through a questionnaire about whether they could perform the test without distraction. Our results show that participants were able to perform the tests in a distraction-free context, given that they reported no distraction in 92% of test sessions and on average high concentration levels. In addition, they reported on average medium-to-good subjective task difficulty across memory tests indicating that while the remote cognitive assessments were challenging, they were neither too easy nor too difficult. Finally, the adherence to the mobile tests was quite good, with a maximum of 20% of participants dropping out after 6 tests within a period of at least 12 weeks. Our results, thus, indicate that it is possible to achieve the level of usability that is required to perform a detailed assessment of episodic memory fully remotely and without any supervision in a memory complainer cohort.

The total testing time required to obtain the RDMC (a single run of ORR, CSR and MDT-OS) was ∼45 minutes. As each test consists of two phases, the 45 minutes result from 6 separate 5–10-minute sessions on 3 different days in separate weeks. In principle, all three tests could be obtained within a single day. However, we decided not to enforce the shortest possible acquisition time. Instead, we decided to leverage the opportunities of mobile and unsupervised testing to achieve a more meaningful implementation. To that end we stretched out the assessment over several weeks to enable a more representative sampling of memory performance over time and thereby be less vulnerable to day-to-day performance fluctuations. We used the spaced testing to ease stress for the patients and eliminate potential implementation problems that would lead to worries and complaints by those patients that felt being tested on a bad day. In an additional regimen, we extended this approach and included even more individual test sessions per paradigm with the aim to increase the robustness of the approach which yielded a higher diagnostic accuracy. Thus, in either scenario, the RDMC reflects memory performance over a period of several weeks rather than a single day, something that would be very difficult to implement with a supervised testing approach. Episodic memory tests such as the FCSRT ^48^ and the other elements of the PACC5 place heavy demands on verbal abilities. This significantly challenges applicability in international trials or in conditions with mild language disorders (e.g., due to a vascular event or primary progressive aphasia) ^4^. The three tests of the RDMC established here, however, are not dependent on verbal abilities such as naming, word-finding or pronunciation and thereby facilitate testing across different dementia syndromes and subtypes of AD as well as in international comparisons. Furthermore, the RDMC shows no overlap with the PACC5 in terms of the paradigm and modalities tested so that there would be no interference with a memory-clinic or trial-based PACC5 or related neuropsychological assessment following case-finding.

In the currently used implementation of the ORR and CSR tests, we did not strictly reinforce adherence to the planned retrieval-delay intervals of these tests, which led some individuals to perform recall and recognition assessments after longer than planned delays. Given that the length of the delay period is associated with task performance, we excluded sessions with significantly extended delay periods ^10^. However, this led to a significant exclusion of participants in our study. Thus, in future implementations of the RDMC, it will be beneficial to optimize usability aspects to a stricter reinforcement of delay intervals.

This study has a number of shortcomings. First, while our results are based on three cohorts from two different countries, this represents a single study with a modest sample size and thus needs to be cross-validated across independent cohorts. Second, while we could show evidence for limited relationships between the RDMC and sample demographics, a large and diverse normative sample is needed in order to calculate demographically-adjusted scores for various covariates. Finally, our sample size was not yet sufficient to assess the relationship with AD biomarkers and the diagnostic accuracy of biomarker stratified subgroups.

Taken together, the high construct validity and retest reliability of the RDMC score paves the way for implementing mobile app-based remote assessment in clinical studies as well as in health care. The current data indicate that the RDMC can facilitate case-finding whenever the main question is about an individual’s MCI-grade cognitive impairment. Future studies need to show whether repeated assessments of the RDMC over time will be sensitive to cognitive change.

## Data Availability

The data, which support this study, are not publicly available, but may be provided upon reasonable request to the authors and pending a material transfer agreement.

## Acknowledgments

We want to thank all participants for their participation in the study.

## Author Contribution

DB and ED were involved in the conceptualization of the study and the preparation of the original draft. IH, MHN, OVB and ND were involved in quality assurance of the remote data collection. DB curated and analyzed the data. WG, LC, KB, MB, AS, KB, RP, AS, ST, JW, SJ, MW, FJ and ED were involved in data collection and participant recruitment. XG and JG contributed analysis tools. All authors reviewed and edited the manuscript.

